# Quantifying population contact patterns in the United States during the COVID-19 pandemic

**DOI:** 10.1101/2020.04.13.20064014

**Authors:** Dennis M. Feehan, Ayesha S. Mahmud

## Abstract

SARS-CoV-2 is transmitted primarily through close, person-to-person interactions. In the absence of a vaccine, interventions focused on physical distancing have been widely used to reduce community transmission. These physical distancing policies can only control the spread of SARS-CoV-2 if they are able to reduce the amount of close interpersonal contact in a population. To quantify the impact of these policies over the first months of the COVID-19 pandemic in the United States, we conducted three waves of contact surveys between March 22 and June 23, 2020. We find that rates of interpersonal contact have been dramatically reduced at all ages in the US, with an 82% (95% CI:80% - 83%) reduction in the average number of daily contacts observed during the first wave compared to pre-pandemic levels. We find that this decline reduced the reproduction number, *R*_0_, to below one in March and early April (0.66, 95% CI:0.35 - 0.88). However, with easing of physical distancing measures, we find increases in interpersonal contact rates over the subsequent two waves, pushing *R*_0_ above 1. We also find significant differences in numbers of reported contacts by age, gender, race and ethnicity. Certain demographic groups, including people under 45, males, and Black and Hispanic respondents, have significantly higher contact rates than the rest of the population. Tracking changes in interpersonal contact patterns can provide rapid assessments of the impact of physical distancing policies over the course of the pandemic and help identify at-risk populations.

## Main

The dynamics of COVID-19 in a population are fundamentally dependent on rates of interpersonal interaction and on patterns of who interacts with whom. With the sharp increase in COVID-19 cases globally, many countries adopted physical distancing practices at an unprecedented scale in an effort to reduce transmission.^1^ On March 16, 2020, seven counties in the San Francisco Bay Area ordered residents to shelter in place in response to evidence of community transmission of COVID-19. Over the subsequent days and weeks, other US cities and states followed suit. At the start of April 2020, the majority of people living in the US were under orders to dramatically restrict their daily activities. By the end of April, however, some localities began easing restrictions, and there is presently considerable heterogeneity in physical distancing policies across US states, counties, and cities (Mervosh, Lu, and Swales 2020).

Strong physical distancing measures are effective in controlling the spread of the virus only if they are able to reduce the amount of close interpersonal contact in a population. To quantify how much interpersonal contact is changing as the pandemic evolves in the US, we developed the Berkeley Interpersonal Contact Survey (BICS). The BICS study collects information about the total number of contacts people have, as well as detailed information about who people are interacting with. This detailed information is particularly important for informing epidemiological models and for identifying populations at greatest risk to COVID-19. Age-structured contact rates are especially relevant for COVID-19 because of age-related variation in clinical outcomes, and possibly susceptibility and transmissibility (Davies et al. 2020).

Here, we describe changes in contact rates and patterns over the course of the pandemic, and identify important correlates of close interpersonal contact in the US. We also evaluate the effectiveness of physical distancing policies by estimating the impact of reduced contact rates on the reproduction number, *R*_0_ – the average number of secondary infections arising from a single infection in a fully susceptible population.

Data collection took place in three waves: between March 22 and April 8, 2020 (pilot study, Wave 0); between April 10 and May 4, 2020 (Wave 1); and between June 17 and 23, 2020 (Wave 2). We surveyed a total of 6,495 respondents in the U.S. (Wave 0 n=1,437, Wave 1 n=2,627, Wave 2 n=2,431). Survey respondents were asked to report the number of people they had contact with on the day before the interview. Respondents reported a total of 27,570 contacts and provided detailed reports about 18,478 contacts. We oversampled respondents in certain cities; analyses here are weighted to account for sample composition (Methods).

Since physical distancing policies are intended primarily to reduce non-household contacts, we investigate both the total number of reported contacts and the number of reported non-household contacts. Figure 1, Panels A-B show histograms of the number of contacts (Panel A) and non-household contacts (Panel B) reported by respondents in each wave. Respondents reported a median of 2 contacts (0 non-household) in Wave 0, a median of 3 contacts (1 non-household) in Wave 1, and a median of 3 contacts (1 non-household) in Wave 2. Qualitatively, the pattern of contacts is similar in each wave, but with higher levels of contact in Wave 1 and Wave 2, when compared to Wave 0. We confirm this increase in contact levels over time with a model-based analysis below.

**Figure 1:**
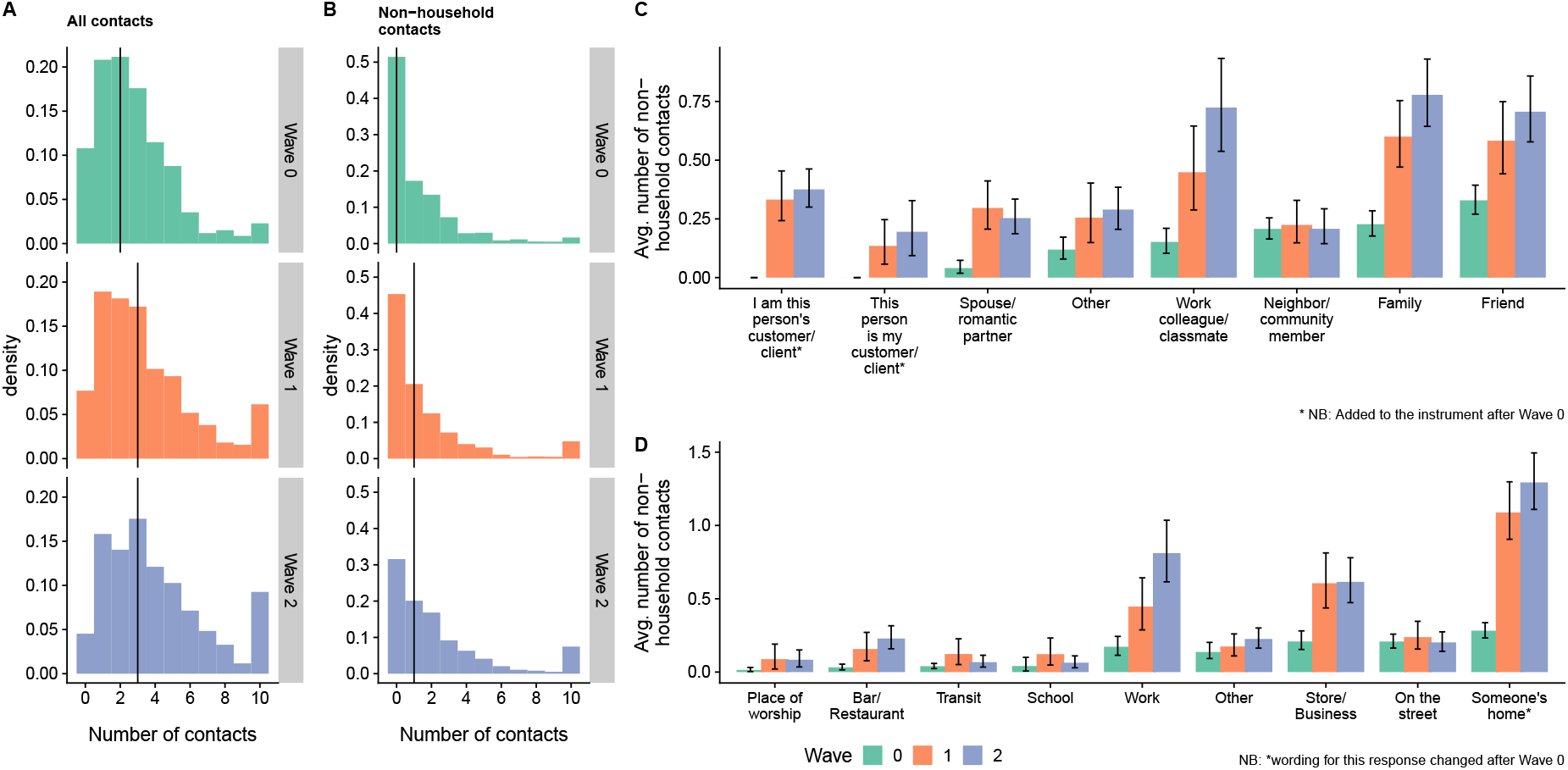
(**A-B**) Histograms of reported number of contacts (A) and non-household contacts (B) among respondents for each wave. Each bar shows estimated average numbers of contacts in each category, per person. For example, the top panel shows that the average respondent reported almost 0.8 contacts to family members in Wave 2. Reported contacts are top-coded at 10 in these plots. (**C-D**) Estimated average number of contacts each person reported to have taken place by contact’s relationship (C) and location (D). Uncertainty estimates are 95% intervals derived from the bootstrap.

For up to three contacts, respondents were asked to report detailed information including the contact’s age, sex, relationship to the respondent, and the location of the contact event. Using this information, we estimated the composition of respondents’ contacts by relationship and by location (Methods). Figure 1 shows the estimated average number of non-household contacts each person reported to have taken place by contact’s relationship (top panel) and location (bottom panel). Across Waves 0, 1, and 2, the average number of interactions with family, friends, and work colleagues increases, and in Waves 1 and 2, these three relationships are responsible for most non-household interpersonal contact. In Wave 0, with contact levels uniformly very low, no one relationship stands out as explaining most non-household interaction.

Across all three waves, the most common location of reported contacts was someone’s home. Across Waves 0, 1, and 2 we find increases in the number of work contacts and home contacts, and between Waves 0 and 1 we see increases in contacts at stores and businesses.

Previous studies have found that during non-pandemic periods the average number of contacts is related to characteristics of people - e.g., age and household size - and to structural factors like day of the week - weekday versus weekend (Mossong et al. 2008). To investigate correlates of contacts in the US during the emergence of COVID-19, we fit negative binomial regression models to data for all contacts and for non-household contacts (see Methods). Figure 2 compares estimated coefficients from these models; coefficient estimates greater (less) than 0 imply an association with higher (lower) reported levels of contact, controlling for other predictors in the model. The model estimates confirm that the level of contact increased from Wave 0 to Wave 1, and again from Wave 1 to Wave 2. Figure 2 also shows that, controlling for other covariates, certain groups of people are estimated to have elevated contact rates, including people under 45; males; and Black and Hispanic respondents. People in the New York and in Boston are estimated to have lower contact rates, net of other predictors in the model.

**Figure 2:**
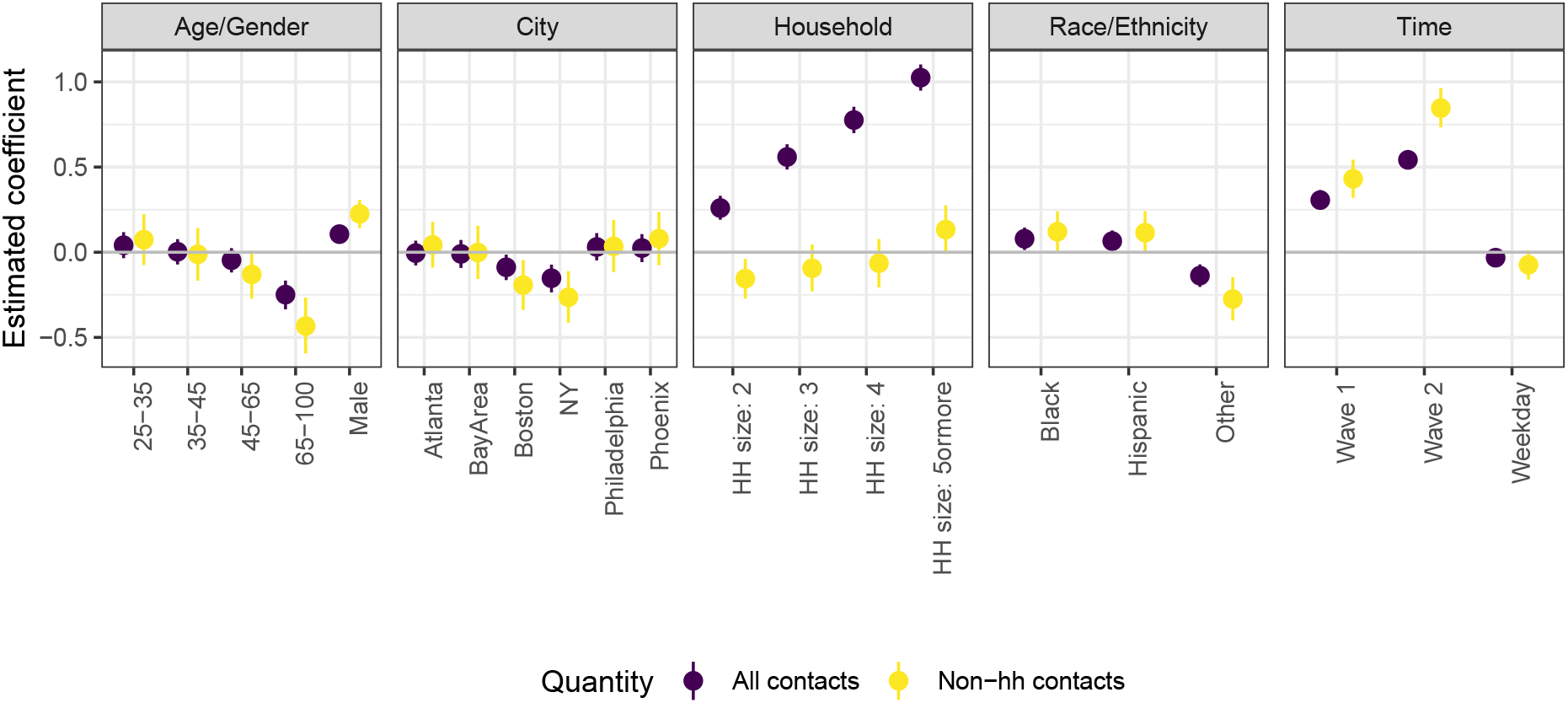
Estimated posterior mean and 95% credible intervals for coefficients from negative binomial models fit to all reported contacts (purple) and to non-household contacts (yellow).

To estimate relative changes in transmission over the course of the pandemic, we estimated the impact of changing contact rates on the reproduction number. According to the *social contact hypothesis*, for respiratory pathogens such as SARS-COV-2, relative changes in *R*_0_, can be estimated by comparing the dominant eigenvalues of age-structured contact matrices (Wallinga, Teunis, and Kretzschmar 2006; Jarvis et al. 2020). We calculated age-structured contact matrices, adjusting for the age distribution of survey respondents and the reciprocal nature of contacts, for each wave of the BICS study. We compare these with baseline data on pre-pandemic contact patterns in the US to understand the impact of physical distancing policies and the implications for the transmission of SARS-CoV-2. There are surprisingly few existing estimates for the rate of contact in the US before the COVID-19 pandemic. We compare our estimates to contact patterns estimated from a probability sample of US Facebook users in 2015 (Feehan and Cobb 2019).

We find large declines in daily interpersonal interaction compared to business as usual, with the largest decline in Wave 0 (82%) followed by Wave 1 (74 %) and Wave 2 (68%). Figure 3 shows the estimated age-structured contact matrix and the reduction in interpersonal contact in each age category for the three BICS waves compared to the 2015 study. We find considerable declines across all age groups, with largest absolute decline in the 25-35 age group. However, even at these low absolute levels of interpersonal contact, we continue to find distinctive patterns of assortative mixing by age found in previous contact studies.

**Figure 3:**
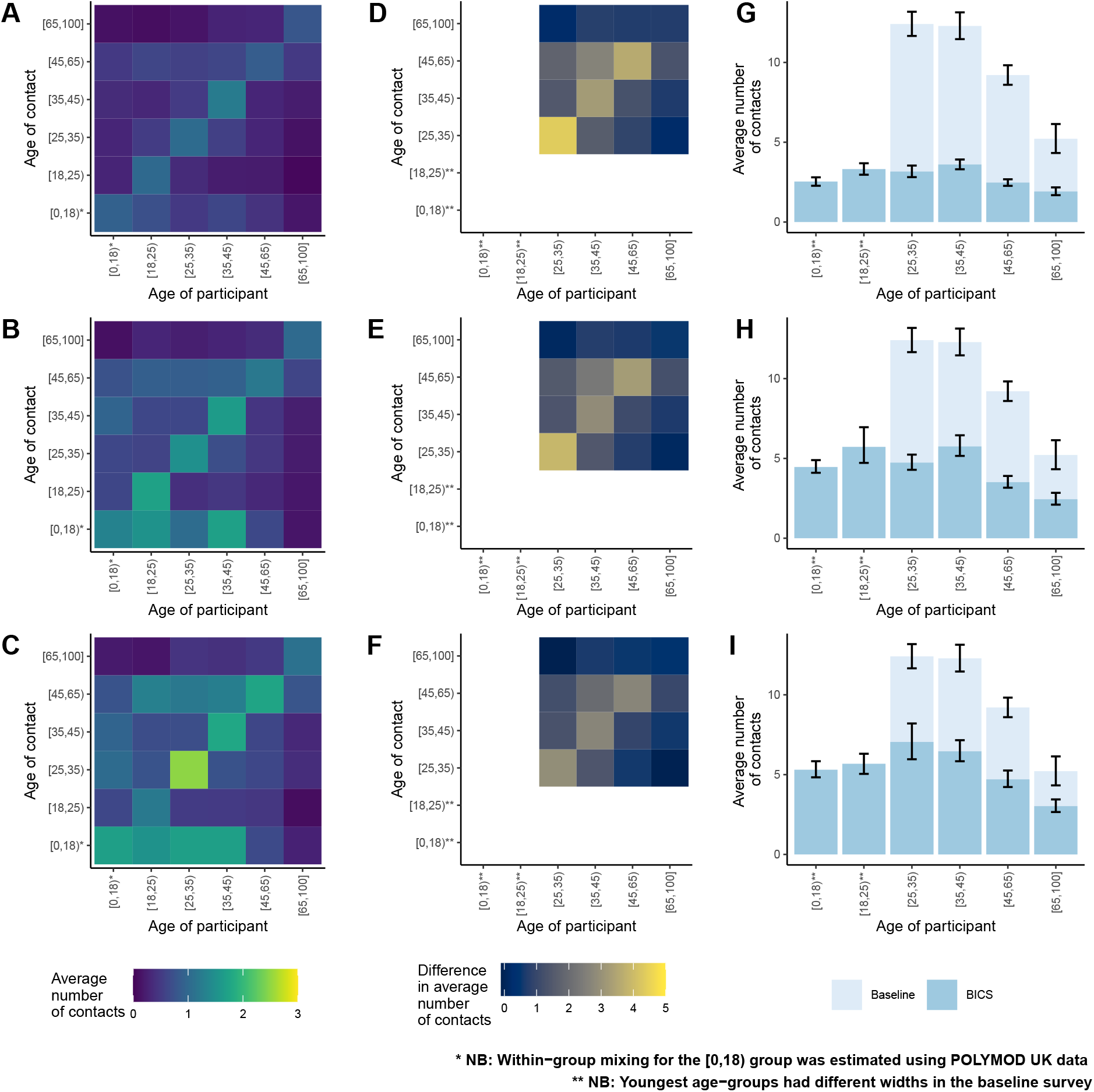
(**A-C**) Age-structured contact matrices from the three BICS waves after adjusting for the age distribution of survey respondents and the reciprocal nature of contacts; lighter colors indicate higher number of average daily contacts. (**D-F**) Difference in the average number of contacts between the 2015 study and the three BICS waves; lighter colors indicate a larger absolute difference between the 2015 study and the BICS data. (G-I) Average number of reported contacts for each respondent age-group for the BICS data (darker color) compared to the 2015 study (lighter color), along with 95% confidence intervals derived from the bootstrap. Top panel shows BICS Wave 0; middle panel shows BICS Wave 1; bottom panel shows BICS Wave 2.

We estimated the relative reduction in *R*_0_, assuming (1) that contact patterns in the population before physical distancing became widespread were equivalent to the 2015 study (Feehan and Cobb 2019), and (2) that disease-specific parameters remained unchanged over the course of the survey period (Methods). We find 73% (95% CI:72 - 75%), 57% (95% CI: 53 - 61%), and 48% (95% CI:43 - 53%) declines in the implied *R*_0_ in Waves 0, 1, and 2 respectively, relative to the pre-pandemic period. The contact patterns observed in our survey suggest a substantial reduction in *R*_0_ under physical distancing, particularly during the Wave 0 study period. Figure 4 shows the *R*_0_ estimates for the three survey waves, assuming an average *R*_0_ value of 2.5 in the absence of physical distancing. The dramatic reduction in contact rates observed in Wave 0 was sufficient in reducing *R*_0_ to 0.66 (95% CI:0.38 - 0.95) in Wave 0. However, with the easing of physical distancing and increase in overall contact rates, *R*_0_ increased to 1.06 (95% CI: 0.61 - 1.54) by Wave 1 and 1.29 (95% CI: 0.73 - 1.85) by Wave 2. We repeat the analysis using contact patterns from UK participants in the POLYMOD study (Mossong et al. 2008), which has been the gold-standard for modeling age-specific contact patterns in many settings, as the pre-pandemic baseline; our results are qualitatively similar (Figure 4).

**Figure 4:**
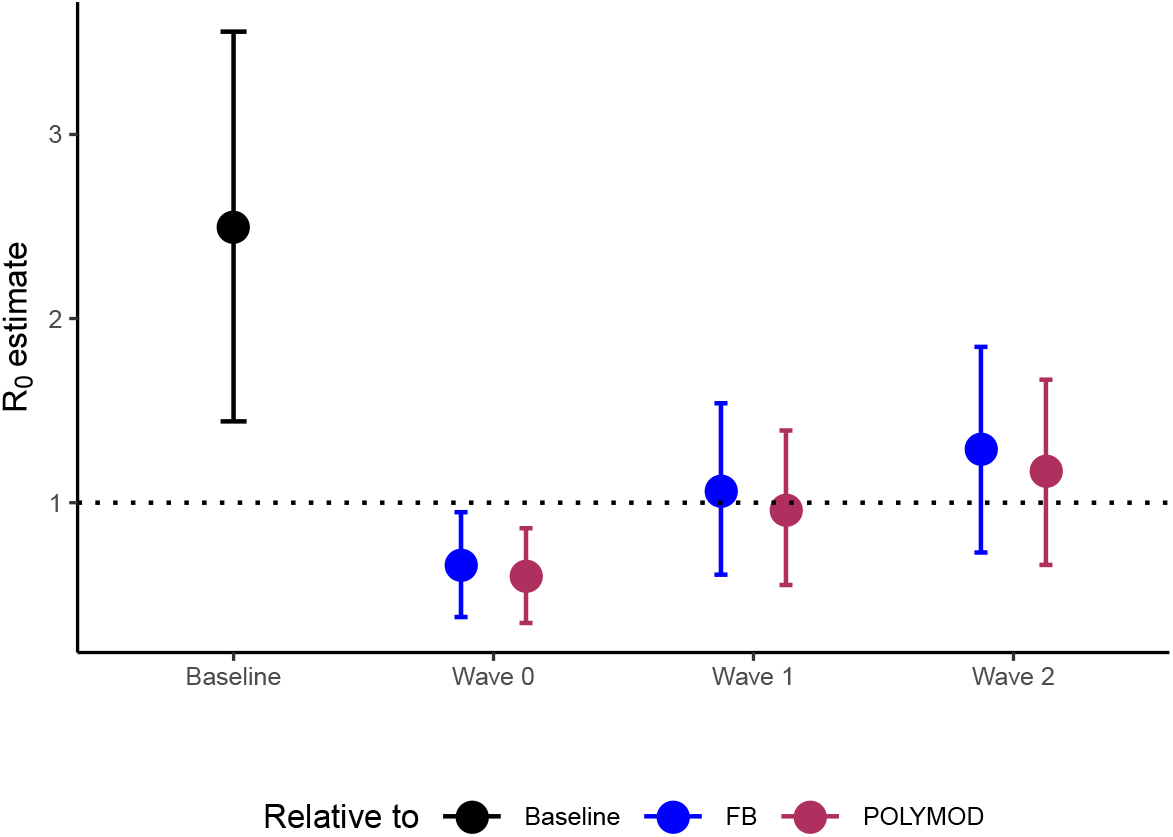
*R*_0_ estimates from the BICS contact matrices for each wave relative to two baseline contact matrices from the 2015 study and the UK POLYMOD study, and assuming a baseline *R*_0_ value drawn from a normal distribution with mean 2.5 and standard deviation of 0.54

**Figure 5:**
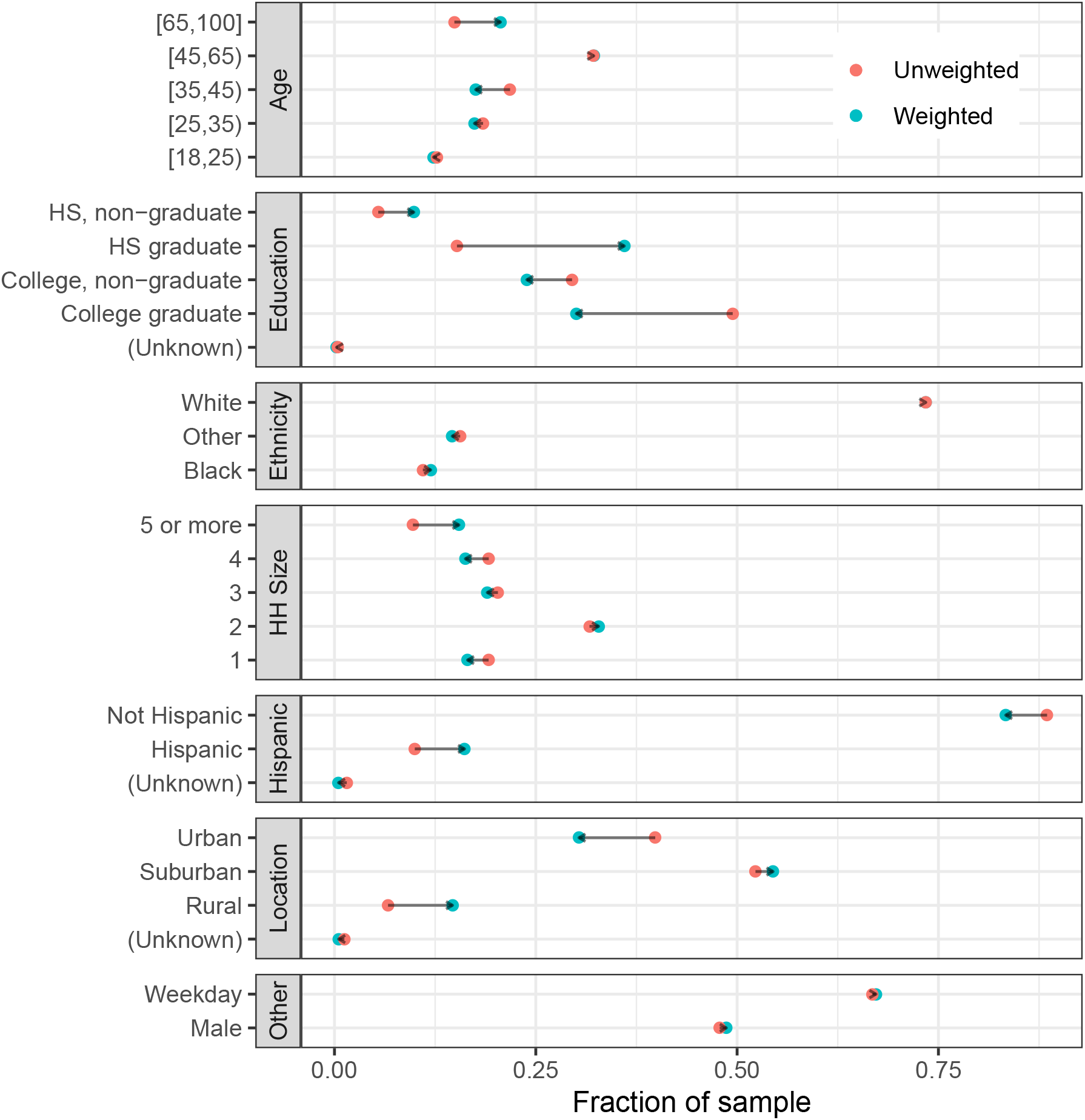
Characteristics of respondents to our survey. We use calibration weights to improve the representativeness of our sample.

We find large reductions in the number of contacts reported in our survey compared to business as usual, suggesting that the physical distancing measures adopted in the U.S. in March and April had their intended impact. Compared to the contact survey conducted in 2015 (Feehan and Cobb 2019), our estimates suggest that in Wave 0 there was about 82% (95% CI: 80% - 83%) reduction in the daily average number of contacts per person. This finding is similar to the declines recently observed in the UK and China (Jarvis et al. 2020; Zhang et al. 2020). As time elapsed, physical distancing policies were relaxed and then, in some jurisdictions, reimposed. We find that over this time period the rate of close interpersonal contacts in the US gradually increased from an unprecedented low level in March, pushing the estimated *R*_0_ values above 1 by June. In addition to an overall increase in the average number of reported contacts, we also find an increase in the number of contacts at work, as well as at stores and businesses; this has implications for SARS-CoV-2 transmission as the economy reopens.

Our analysis here has several important limitations. In this study, we used a quota sample from an online panel rather than a probability sample. Previous contact studies have also used various alternatives to probability samples (e.g., Eames et al. 2012, 2010; Grijalva et al. 2015; Ibuka et al. 2016; Klepac, Kissler, and Gog 2018). Online panels allow data to be collected rapidly and frequently, whereas the time and cost required to design and implement a probability sample are prohibitive. Further, obtaining a probability sample during a pandemic is further complicated by the logistical challenges arising from the need to protect interviewers and respondents. However, future work based on a national probability sample would be a valuable complement to our study.

There may be some recall bias in our survey estimates, as respondents were asked to report on contacts from the previous day. There may also be social desirability bias arising from awareness of social distancing policies. Our surveys were only conducted in English, meaning that we are not able to reach people who only speak other languages. We do not survey children, and are unable to capture contacts within age groups below the age of 18. Finally, our estimates of relative changes in *R*_0_ do not take into account possible age-specific differences in susceptibility or infectiousness, or changes in infection transmissibility due to other factors such as the use of face masks.

The BICS study is ongoing, and will continue to collect data for the next several months, with the goal of measuring changes in contact patterns as interventions change and schools and workplaces reopen. The data from the BICS study provides a unique opportunity to understand how interpersonal contact patterns are changing in the US over the course of the pandemic, and the epidemiological implications for COVID-19 and other respiratory pathogens. Future work will focus on applying these estimates to parameterize age-structured mathematical models of SARS-CoV-2 transmission and to monitor and evaluate the effectiveness of physical distancing policies over time.

## Data Availability

Our dashboard has the estimated mixing matrices, and we intend to provide a public-use version of the survey microdata as soon as possible.

https://contact-survey.github.io/dashboard/

## A Methods

### Survey methodology

We designed and fielded a survey to measure interpersonal interaction in the United States. Following the POLYMOD project (Mossong et al. 2008) and subsequent studies (e.g., Zhang et al. 2020; Jarvis et al. 2020; Dorélien et al. n.d.), survey respondents were asked to report the number of people they had conversational contact with on the day before the interview; in Waves 1 and 2, we also asked about physical contact. Respondents were asked to provide detailed information about up to three of their reported contacts; this detailed information included who those contacts were, how long those contacts lasted, and where they took place. In Wave 0, respondents were asked to report all contacts, and to then report how many of their contacts were not household members. In Waves 1 and 2, respondents were asked to provide a household roster, and then report only contacts outside of the household.

Respondents were recruited using Lucid, an online panel provider. In each wave, we obtained two samples: first, a quota sample that is intended to be representative of the United States; and, second, several smaller quota samples from specific cities: New York, the San Francisco Bay Area, Atlanta, Phoenix, and Boston. In Wave 1, Philadelphia was added.

The project was approved by the UC Berkeley IRB (Protocol 2020-03-13128).

### Weighting

#### Respondent-level weights

We adopt a model-based approach to inference, which is appropriate for our quota sample (Elliott and Valliant 2017). Except where noted, we pool results from the national and city samples together in this analysis. We use calibration to produce pseudo-probabilities of inclusion, and use these pseudo-probabilities of inclusion as the basis for weights used to make population-level inferences (Deville and Särndal 1992; Särndal and Lundstrôm 2005). We calibrate based on: age categories (18-23, 24-29, 30-39, 40-49, 50-59, 60-69, 70+); sex; age by sex interactions; education (non-high school graduate, high school graduate, some college, college graduate); race (white, black, other); Hispanicity; household size category (1, 2, 3, 4, 5 or more); and whether the respondent’s county is rural/suburban/urban. All population values except for rural/suburban/urban are taken from a 1-year extract of the 2018 American Community Survey provided by IPUMS (Ruggles et al. 2020). We ascertain whether each respondent lives in an urban, suburban, or rural area by mapping the respondent’s zip code to county, and then using the [county-level urban/suburban/rural codes from the CDC](https://www.cdc.gov/nchs/data_access/urban_rural.htm). In order to map zip code to county, we use the crosswalk developed by Sood (Sood 2016). We perform the calibration using the R packages autumn (https://github.com/aaronrudkin/autumn) and leafpeepr (https://rdrr.io/github/rossellhayes/leafpeepr/).

#### Contact-level weights

In Wave 0, the pilot study, respondents were asked for their total number of contacts and for the number of contacts who were not household members. Then, respondents were asked to provide detailed information for three of these contacts; this detailed information included contact age, sex, relationship to respondent, and contact location. If respondents reported more than three total contacts, they were asked to report in detail about the first three contacts who came to mind. In Waves 1 and 2, respondents were asked to report about the age and sex of all of their household members, and then to report the number of contacts they had with non-household members. Respondents were then asked toreport detailed information for the first three non-household member contacts who came to mind.

In all waves, some respondents reported more than 3 total contacts, but only provided detailed information about three contacts. In these cases, in order to make inferences about the total number of contacts, we use within-respondent weights. For example, suppose respondent *i* reports a total of *d_i_* = 6 contacts, and provides detailed information about 3 of them. Then each of the three contacts receives a weight of 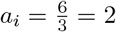. If, on the other hand, respondent *j* reports a total of *d_j_ =* 2 contacts and provides detailed information about both of them, then *a_j_ =* 1. Conceptually, *a_i_* is the number of respondent *i*’s contacts represented by each contact who gets reported about in detail. Feehan and Cobb (2019) discusses this weighting approach in greater detail.

When we make population level inferences about contact characteristics, such as the relationship and location distributions shown in Figure 1, we use these contact weights in combination with the respondent weights. For example, to estimate the proportion of contacts at work, we use

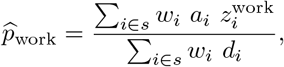

where

- *s* is the sample of all respondents
- *w_i_* is the respondent-level calibration weight
- *a_i_* is the within-respondent weight for respondent *i*’s contacts
- 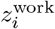 is a variable that has how many of respondent *i*’s detailed contacts were reported to have happened at work
- *d_i_* is the total number of contacts respondent *i* reports

The intuition is that *w_i_* is the number of people respondent *i* represents in the general population, and *a_i_* is the number of *i*’s contacts that is represented by each detailed contact. Feehan and Cobb (2019) has additional details about this approach.

### City samples

For our city-specific samples, we recruited respondents who lived in the Designated Market Area (DMA) surrounding each city. DMAs are intended to capture media markets, and therefore often include much more than just the urban core of a city.

Our city-specific samples recruited respondents in the DMAs associated with six cities: Atlanta, the San Francisco Bay Area, Boston, New York, Phoenix and, starting in Wave 1, Philadelphia. Figure 6 shows the media markets.

**Figure 6:**
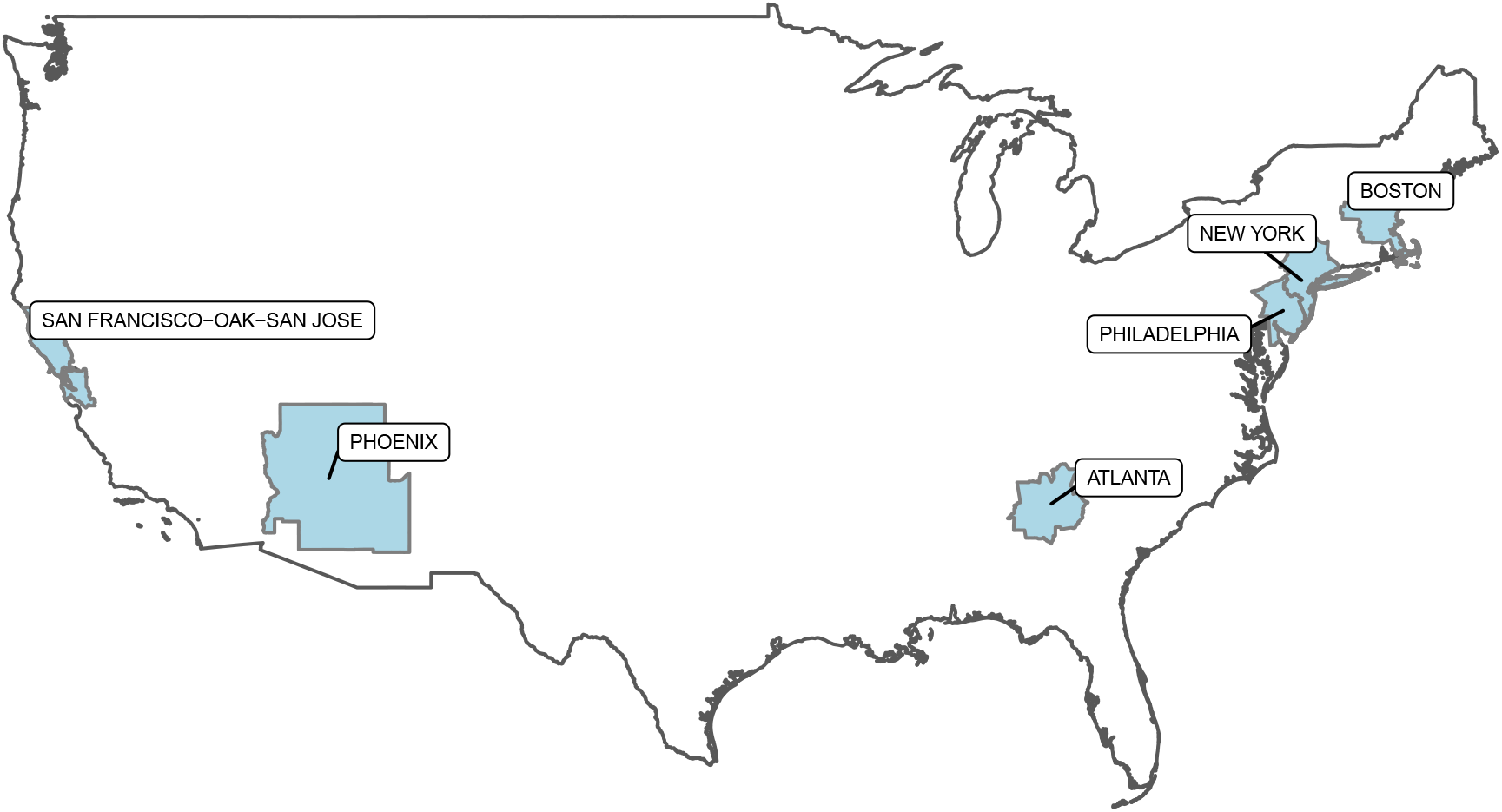
The geographical areas corresponding to the cities in our sample.

### Statistical Model

To investigate factors associated with interpersonal contacts, we developed statistical models. We fit separate models to the total reported contacts and to the number of reported non-household contacts. In each case, we model the expected number of contacts using a negative-binomial distribution. The negative binomial distribution is appealing because it allows for overdispersion – that is, it enables us to model count data that exhibit more variance than would be expected under a Poisson distribution. This modeling approach has previously been used to study contact data (e.g., Mossong et al. 2008).

In our model, the log of the expected number of contacts for respondent *i* is given by

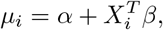

where *X_i_* is a vector of covariates that includes age category, gender, household size, survey wave, city, ethnicity (Black / White/ Other), Hispanicity, and whether or not the day being reported about is a weekday. *β* is a vector of coefficients to be estimated.

Given *μ_i_*, we define *λ_i_ =* exp(*μ_i_*) to be the expected number of contacts for respondent *i*. Then we model the reported number of contacts for respondent *i*, *y_i_*, as

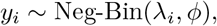

where *ϕ* ∈ [1, *∞*) is a shape parameter that is inversely related to overdispersion; that is, the higher *ϕ* is estimated to be, the more similar *y*_i_’s distribution is to a Poisson distribution with rate parameter *λ_i_*.

In our data, observations from Wave 0 are censored above 10, because the survey instrument allowed respondents to report up to ‘10 or more’ contacts. Wave 1 and Wave 2 allowed respondents to enter any number of contacts, but we topcoded contacts at 29, following Mossong et al. (2008). Reports that are topcoded or censored in any of the three waves are treated as right-censored in the model. We adopt a Bayesian approach to fitting the model. For all of the regression coefficients *β*, we assume flat priors. For the intercept and the shape parameter, we assume very weak priors. Specifically, we assume a priori that the intercept *α* is distributed with mean 0 and a large variance by using pr(*α*) ~ Student-t(3, 0,10); And we assume a priori that the the shape parameter *ϕ*, is distributed with mean 1 and a very large variance by using pr(*ϕ*) ~ Gamma(.01, .01). Figure 2 shows posterior estimates for the mean and 95% credible intervals for key coefficients of interest. Table 1, shows summary statistics for the predictors used in our model.

**Table 1:** Characteristics of the sample.

|  | Unweighted |  |  |  | Weighted |  |  |  |
| --- | --- | --- | --- | --- | --- | --- | --- | --- |
|  | Overall, N = 6495 | Wave 0, N = 1,437 | Wave 1, N = 2,627 | Wave 2, N = 2,431 | Overall, N = 6495 | Wave 0, N = 1,437 | Wave 1, N = 2,627 | Wave 2, N = 2,431 |
| **Gender** |  |  |  |  |  |  |  |  |
| **Female** | 3,388 (52%) | 756 (53%) | 1,376 (52%) | 1,256 (52%) | 3,334 (51%) | 738 (51%) | 1,348 (51%) | 1,248 (51%) |
| **Male** | 3,107 (48%) | 681 (47%) | 1,251 (48%) | 1,175 (48%) | 3,161 (49%) | 699 (49%) | 1,279 (49%) | 1,183 (49%) |
| **Age** |  |  |  |  |  |  |  |  |
| **[18,25)** | 827 (13%) | 187 (13%) | 346 (13%) | 294 (12%) | 798 (12%) | 170 (12%) | 339 (13%) | 289 (12%) |
| **[25,35)** | 1,198 (18%) | 280 (19%) | 469 (18%) | 449 (18%) | 1,128 (17%) | 277 (19%) | 445 (17%) | 406 (17%) |
| **[35,45)** | 1,416 (22%) | 300 (21%) | 557 (21%) | 559 (23%) | 1,138 (18%) | 242 (17%) | 451 (17%) | 445 (18%) |
| **[45,65)** | 2,087 (32%) | 456 (32%) | 861 (33%) | 770 (32%) | 2,092 (32%) | 466 (32%) | 865 (33%) | 760 (31%) |
| **[65,100]** | 967 (15%) | 214 (15%) | 394 (15%) | 359 (15%) | 1,339 (21%) | 281 (20%) | 528 (20%) | 530 (22%) |
| **City** |  |  |  |  |  |  |  |  |
| **National** | 3,163 (49%) | 644 (45%) | 1,392 (53%) | 1,127 (46%) | 3,385 (52%) | 685 (48%) | 1,528 (58%) | 1,171 (48%) |
| **Atlanta** | 628 (9.7%) | 210 (15%) | 203 (7.7%) | 215 (8.8%) | 644 (9.9%) | 217 (15%) | 191 (7.3%) | 236 (9.7%) |
| **Bay Area** | 564 (8.7%) | 133 (9.3%) | 206 (7.8%) | 225 (9.3%) | 457 (7.0%) | 122 (8.5%) | 151 (5.8%) | 184 (7.6%) |
| **Boston** | 569 (8.8%) | 158 (11%) | 199 (7.6%) | 212 (8.7%) | 562 (8.7%) | 158 (11%) | 188 (7.2%) | 215 (8.9%) |
| **NY** | 587 (9.0%) | 150 (10%) | 213 (8.1%) | 224 (9.2%) | 506 (7.8%) | 131 (9.1%) | 189 (7.2%) | 185 (7.6%) |
| **Philadelphia** | 435 (6.7%) | 0 (0%) | 219 (8.3%) | 216 (8.9%) | 474 (7.3%) | 0 (0%) | 226 (8.6%) | 249 (10%) |
| **Phoenix** | 549 (8.5%) | 142 (9.9%) | 195 (7.4%) | 212 (8.7%) | 467 (7.2%) | 124 (8.7%) | 153 (5.8%) | 190 (7.8%) |
| **Urban/Rural** |  |  |  |  |  |  |  |  |
| **Urban** | 2,587 (40%) | 578 (41%) | 1,021 (39%) | 988 (41%) | 1,972 (31%) | 436 (31%) | 798 (31%) | 738 (31%) |
| **Suburban** | 3,396 (53%) | 733 (52%) | 1,409 (54%) | 1,254 (52%) | 3,538 (55%) | 783 (55%) | 1,431 (55%) | 1,324 (55%) |
| **Rural** | 432 (6.7%) | 104 (7.3%) | 169 (6.5%) | 159 (6.6%) | 952 (15%) | 211 (15%) | 385 (15%) | 356 (15%) |
| **Ethnicity** |  |  |  |  |  |  |  |  |
| **White** | 4,768 (73%) | 1,044 (73%) | 1,942 (74%) | 1,782 (73%) | 4,770 (73%) | 1,059 (74%) | 1,917 (73%) | 1,794 (74%) |
| **Black** | 713 (11%) | 174 (12%) | 292 (11%) | 247 (10%) | 776 (12%) | 167 (12%) | 308 (12%) | 301 (12%) |
| **Other** | 1,014 (16%) | 219 (15%) | 393 (15%) | 402 (17%) | 949 (15%) | 211 (15%) | 402 (15%) | 337 (14%) |
| **Hispanic** | 648 (10%) | 133 (9.4%) | 273 (11%) | 242 (10%) | 1,049 (16%) | 232 (16%) | 424 (16%) | 393 (16%) |
| **Education** |  |  |  |  |  |  |  |  |
| **College graduate** | 3,212 (50%) | 667 (47%) | 1,300 (50%) | 1,245 (51%) | 1,950 (30%) | 431 (30%) | 789 (30%) | 729 (30%) |
| **Some college** | 1,916 (30%) | 452 (32%) | 813 (31%) | 651 (27%) | 1,551 (24%) | 343 (24%) | 628 (24%) | 580 (24%) |
| **High school graduate** | 988 (15%) | 249 (17%) | 400 (15%) | 339 (14%) | 2,338 (36%) | 517 (36%) | 946 (36%) | 875 (36%) |
| **Non-high school graduate** | 353 (5.5%) | 62 (4.3%) | 106 (4.0%) | 185 (7.6%) | 640 (9.9%) | 142 (9.9%) | 259 (9.9%) | 240 (9.9%) |
| **Household Size** | 2.00 (2.00, 4.00) | 2.00 (2.00, 4.00) | 2.00 (2.00, 4.00) | 2.00 (2.00, 4.00) | 3.00 (2.00, 4.00) | 3.00 (2.00, 4.00) | 3.00 (2.00, 4.00) | 3.00 (2.00, 4.00) |
| **Weekday** | 4,339 (67%) | 702 (49%) | 1,918 (73%) | 1,719 (71%) | 4,368 (67%) | 684 (48%) | 1,916 (73%) | 1,768 (73%) |

<sup>1</sup> Statistics presented: n (%); median (IQR)

Accounting for this censoring, in our models the log posterior of the parameters given the data, logpr(*α*, *β*, *ϕ|y, X*), is proportional to

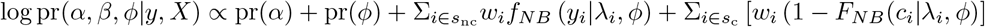

where *s_nc_* is the set of responses that are not censored; *s_c_* is the set of responses that are right-censored, with response *i* ∈ *s_c_* being censored at value *c_i_*; 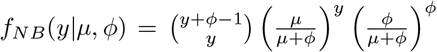 is the PMF of the negative binomial distribution, and *F_NB_* is the cumulative distribution function 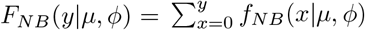; and 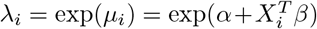 is the expected number of contacts or non-household contacts for respondent *i*. (The parameterizations here are the ones used in stan). For each model, we run 4 chains of the sampler; each chain was run for 1000 warmup iterations and then 1000 sampling iterations. All R-hat statistics are 1, suggesting that the chains mixed effectively.

### Epidemiological Model

We estimate age-structured contact matrices for each wave of the BICS study using previously described methods (Wallinga, Teunis, and Kretzschmar 2006; Mossong et al. 2008; Klepac, Kissler, and Gog 2018; Jarvis et al. 2020). We group respondents and their contacts into six age bins: 0-18, 18-25, 25-35, 35-45, 45-65, 65+. For each age group, we estimate the average daily number of contacts reported by respondents in that age group with contacts in every age group. In other words, our raw contact matrix, M, has entries *m_ij_* which is the average number of daily contacts between respondents in age group, *j*, with their reported contacts in age group, *i*. Adjusting for survey weights, we calculate *m_ij_* as:

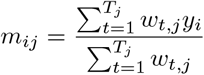

where *w_t,j_* is the weight for particpant, *t*, in age group *j*, and *y_i_* is the weight for their reported contact in age group *i*. *T_j_* is the total number of respondents in age group, *j*.

Contacts in the population must be reciprocial but due to differences in reporting in the survey our raw social contact matrix, M, is not. We impose reciprocity by:

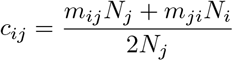

where *c_ij_* are the entries of the reciprocal contact matrix, C, and *N_i_* and *N_j_* the population size in age classs *i* and *j*, respectively. For the youngest age group, for which we have no survey respondents, we assume:

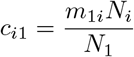

We estimate within age group average number of contacts, *c_ii_*, for the youngest age group using previously developed methods (Klepac, Kissler, and Gog 2018; Jarvis et al. 2020) and data from the United Kingdom POLYMOD study (Mossong et al. 2008). For each wave of the BICS study, we calculate the ratio of the dominant eigenvalue for the contact matrix estimated from the BICS data to the dominant eigenvalue of the contact matrix from the POLYMOD study, with school contacts removed to reflect current school closures, for all age groups that are overlapping between the two studies. The within age group average number of contacts for the [0,18) group in the POLYMOD study is then scaled by this ratio to impute *c*_[0,18)[0,18)_ in the BICS contact matrix.

The transmission dynamics of infectious diseases is summarized by the next generation matrix, **N**, that determines how an infection spreads when a pathogen is first introduced into a fully susceptible population. The basic reproduction number, *R*_0_, is the average number of secondary infections arising from a single infection in a fully susceptible population, and is typically estimated as the spectral radius (dominant eigenvalue), *ρ*(**N**) of the next generation matrix, **N** (Farrington, Kanaan, and Gay 2001). The **N** matrix is proportional to the population contact matrix, **C**. The exact relationship between **N** and **C** is model-dependent, but for respiratory pathogens such as SARS-CoV-2, N is typically modeled as **C** scaled by the duration of infectiousness, 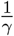, and the probability of transmission for a single contact, *q*. Therefore, the spectral radius of **N**:

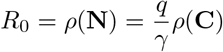

where *ρ*(**C**) is the dominant eigenvalue of the reicprocal population contact matrix. In other words, *R*_0_ is proportional to the dominant eigenvalue of **C**.

Since *R*_0_ is proportional to the dominant eigenvalue of **C**, relative differences in *R*_0_ under different contact patterns is equivalent to the the ratios of the dominant eigenvalues of the different contact matrices. Specifically, if we assume that contact patterns in the population before physical distancing became widespread are equivalent to a baseline contact matrix, and that disease-specific parameters remained unchanged over the course of the survey period, the relative reduction in *R*_0_ during physical distancing, compared to the baseline, is equivalent to the ratios of the dominant eigenvalues of the **C** matrices from the BICS study, **C**^BICS^, to the dominant eigenvalue of the baseline pre-pandemic contact matrix **C**^baseline^:

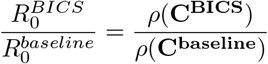

Further, if we assume a distribution for *R*_0_ for COVID-19 in the absence of physical distancing, we can estimate the implied *R*_0_ during the study period, by multiplying this ratio with the *R*_0_ value in the absence of physical distancing. We assume that *R*_0_ prior to physical distancing followed a normal distribution with mean 2.5 and standard deviation of 0.54 based on estimates from literature (Anderson et al. 2020; Jarvis et al. 2020). We compare the BICS contact matrices to two baseline business-as-usual scenarios: contact patterns estimated by (Feehan and Cobb 2019), from a probability sample of US Facebook users, and contact patterns from the UK POLYMOD study (Mossong et al. 2008), that has been widely used in many settings. We compute confidence intervals for the estimated *R*_0_ by repeating the age-imputation and relative *R*_0_ estimation on 5,000 bootstrapped samples from the BICS, POLYMOD and 2015 study contact matrices.

### Bootstraped contact matrices

Uncertainty estimates for the descriptive results - including Figure 1 and the *R*_0_ analysis summarized in Figure 4 - are based on the bootstrap. To obtain bootstrap resamples, we resampled respondents separately in each stratum (i.e., in each city and in the national sample). For each bootstrap resample, we first drew *n_c_* samples with replacement from among the *n_c_* respondents in city c. For resampled respondents who provided detailed reports about all contacts (i.e., those respondents who had *a_i_* = 1 for all contacts *i*), we used the reported detailed contacts without resampling. For resampled respondents who did not provide detailed reports about all contacts, but who reported about a subset of contacts (i.e., for each respondent who had *a_i_ >* 1 for some contact i), we resampled *r_i_* contacts with replacement from among the respondent’s *r_i_* detailed contacts. This second stage is intended to capture sampling variation due to the respondent choosing which contacts to report about from among her total contacts. Using this approach, we obtained 5,000 bootstrap resamples of our dataset, and these bootstrap resamples are the basis for uncertainty estimates.

### Relationships and locations

Here, we present Tables containing the values shown in Figure 1, Panels C and D. These numerical values may be useful as numerical inputs for future modeling work.

| Relationships |  |  |  |
| --- | --- | --- | --- |
|  | Wave 0 | Wave 1 | Wave 2 |
| I am this person’s customer/client* |  | 0.33 (0.24, 0.45) | 0.38 (0.3, 0.46) |
| This person is my customer/client* |  | 0.14 (0.06, 0.25) | 0.2 (0.09, 0.33) |
| Spouse/romantic partner | 0.04 (0.02, 0.07) | 0.3 (0.21, 0.41) | 0.25 (0.19, 0.33) |
| Other | 0.12 (0.08, 0.17) | 0.26 (0.15, 0.4) | 0.29 (0.21, 0.38) |
| Work colleague/classmate | 0.15 (0.1, 0.21) | 0.45 (0.29, 0.65) | 0.72 (0.54, 0.93) |
| Neighbor/community member | 0.21 (0.16, 0.25) | 0.22 (0.15, 0.33) | 0.21 (0.14, 0.29) |
| Family | 0.23 (0.18, 0.28) | 0.6 (0.47, 0.75) | 0.78 (0.64, 0.93) |
| Friend | 0.33 (0.27, 0.39) | 0.58 (0.44, 0.75) | 0.71 (0.58, 0.86) |

\* These relationships were added to the instrument after Wave 0.

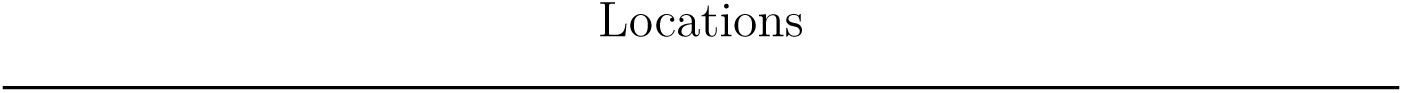
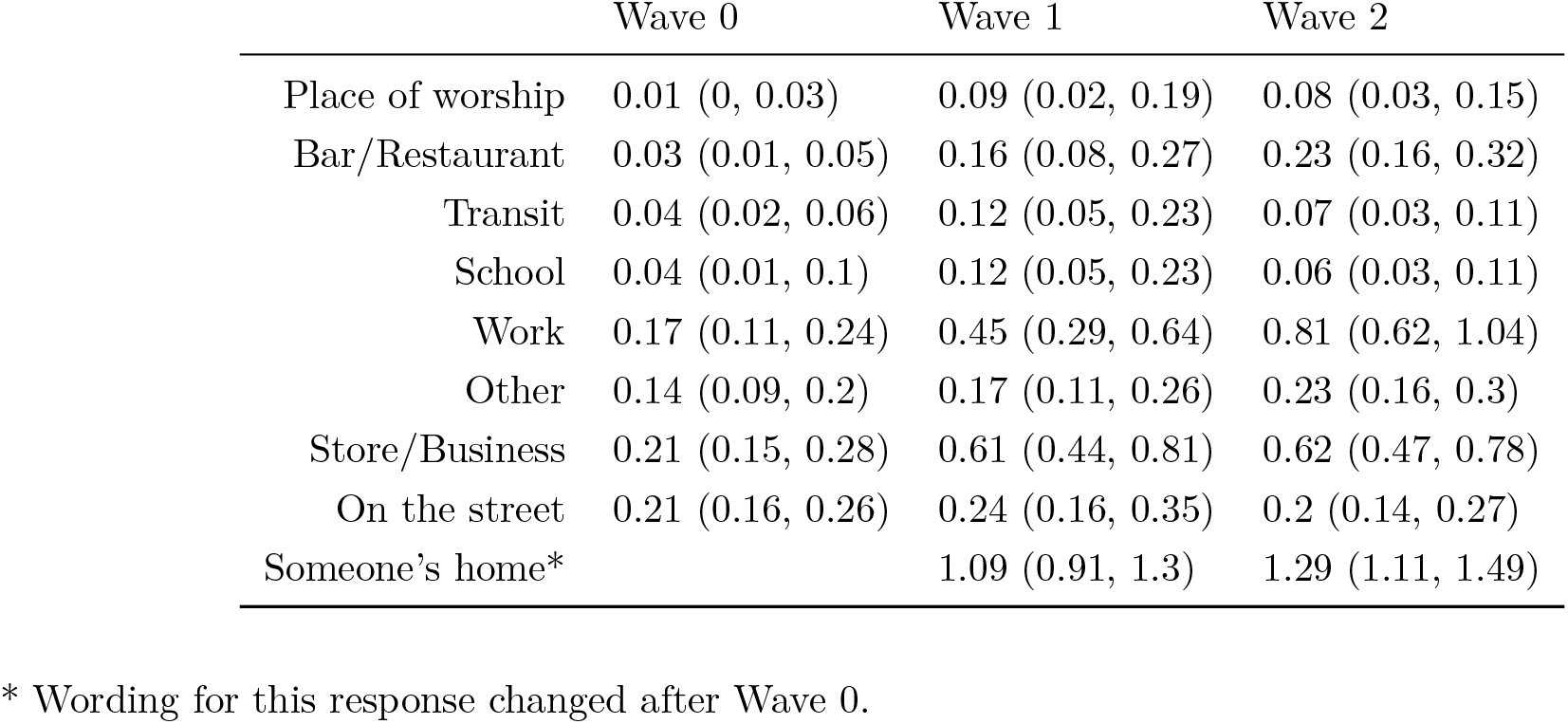

Locations
|  | Wave 0 | Wave 1 | Wave 2 |
| --- | --- | --- | --- |
| Place of worship | 0.01 (0, 0.03) | 0.09 (0.02, 0.19) | 0.08 (0.03, 0.15) |
| Bar/Restaurant | 0.03 (0.01, 0.05) | 0.16 (0.08, 0.27) | 0.23 (0.16, 0.32) |
| Transit | 0.04 (0.02, 0.06) | 0.12 (0.05, 0.23) | 0.07 (0.03, 0.11) |
| School | 0.04 (0.01, 0.1) | 0.12 (0.05, 0.23) | 0.06 (0.03, 0.11) |
| Work | 0.17 (0.11, 0.24) | 0.45 (0.29, 0.64) | 0.81 (0.62, 1.04) |
| Other | 0.14 (0.09, 0.2) | 0.17 (0.11, 0.26) | 0.23 (0.16, 0.3) |
| Store/Business | 0.21 (0.15, 0.28) | 0.61 (0.44, 0.81) | 0.62 (0.47, 0.78) |
| On the street | 0.21 (0.16, 0.26) | 0.24 (0.16, 0.35) | 0.2 (0.14, 0.27) |
| Someone's home* |  | 1.09 (0.91, 1.3) | 1.29 (1.11, 1.49) |

\* Wording for this response changed after Wave 0.

### Sensitivity analysis for contact definition

In Wave 0 (the pilot study), contact was defined using ‘conversational contact’, as explained in this text:

> By **in-person conversational contact**, we mean a two-way conversation with three or more words in the physical presence of another person.
>
> You might have conversational contact with family members, friends, co-workers, store clerks, bus drivers, and so forth.
>
> **(Please do not count people you contacted exclusively by telephone, text, or online**. Only consider people you interacted with face-to-face.)

After the pilot study, in Waves 1 and 2, the survey instrument was modified and contact was defined using this text:

> Now we would like to ask you some questions about people you had **in-person contact** with yesterday.
>
> By **in-person contact**, we mean
>
> **EITHER** a **two-way conversation** with three or more words in the physical presence of another person **OR** physical **skin-to-skin contact** (for example a handshake, hug, kiss, or contact sports)
>
> You might have in-person contact with family members, friends, co-workers, store clerks, bus drivers, and so forth.
>
> **Please do not count people you contacted exclusively by telephone, text, or online**. Only consider people you interacted with face-to-face.

In the main text, for Waves 1 and 2 we combine physical and conversational contacts together, since the combination of these two is most relevant for disease transmission. However, 9 percent of non-household detailed contacts in Waves 1 and 2 were reported to be strictly physical (and not conversational). To assess how sensitive our results are to including all contacts in our analysis, we repeated key analyses excluding these contacts reported to be only physical in Waves 1 and 2. Figures 7 and 8 suggest that this decision does not substantively affect our conclusions.

**Figure 7:**
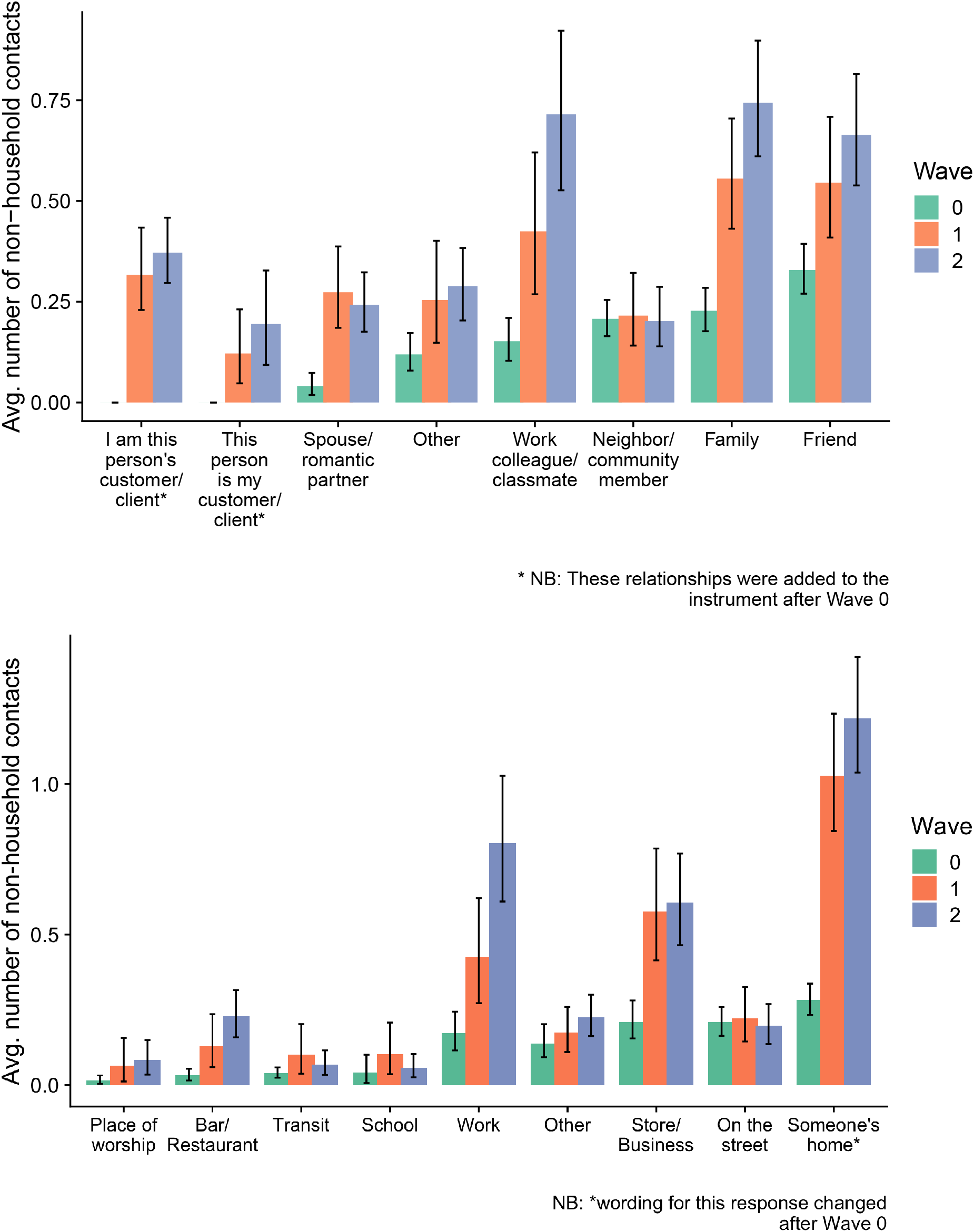
Estimated average number of contacts each person reported to have taken place by contact’s relationship (top panel) and location (bottom panel). Uncertainty estimates are 95% intervals derived from the bootstrap. Contacts are restricted to those who were conversational contacts for Waves 1 and 2.

**Figure 8:**
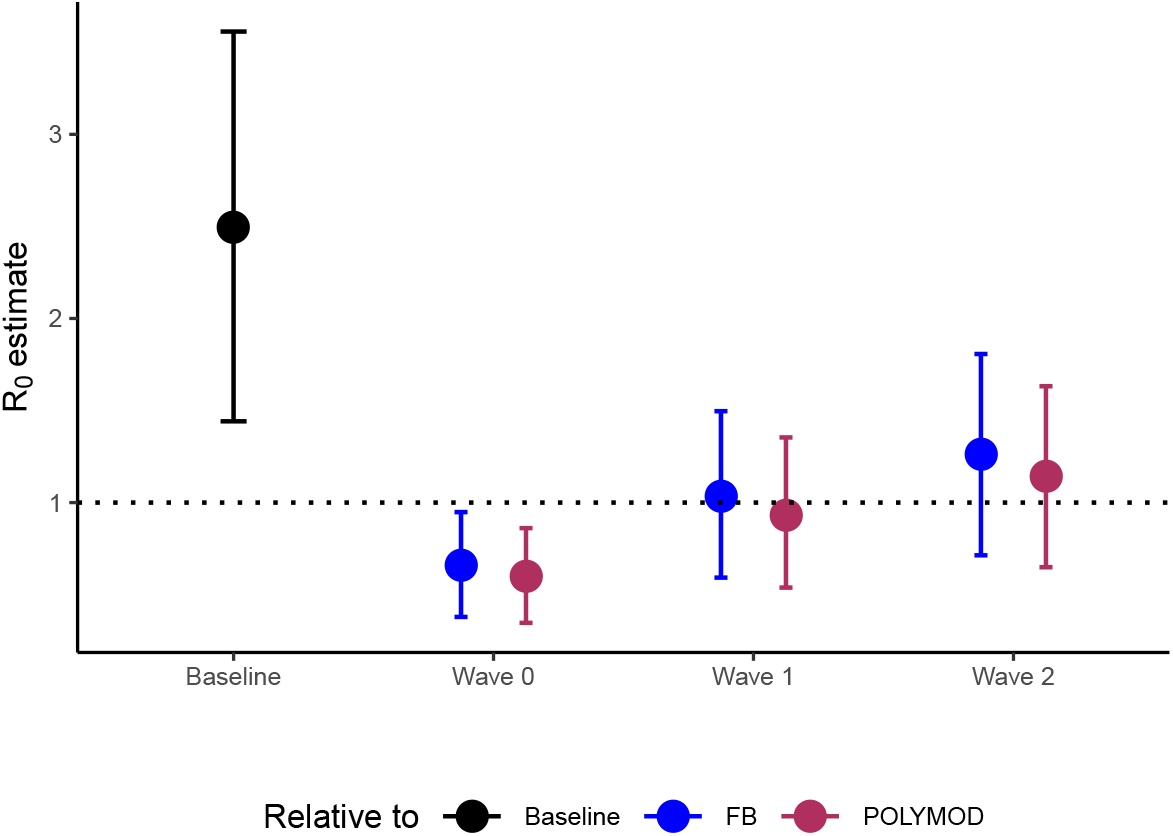
Estimated *R*_0_ for each survey wave. Contacts are restricted to those who were conversational contacts for Waves 1 and 2.

## Footnotes

* For helpful feedback on these results, we thank participants in the April 1, 2020 Berkeley Population Center Brown Bag, C. Jessica E. Metcalf, Caroline Buckee, and Audrey Dorelien. Seed funding was provided by a Berkeley Population Center pilot grant (NICHD P2CHD073964).

1 Throughout the paper, we use the term ‘physical distancing’; in the popular press, this concept is often referred to as ‘social distancing’.

## Notes

### Competing Interest Statement

The authors have declared no competing interest.

### Summary of Updates

We have updated the manuscript with more recent data and additional analyses.

